# An International, Expert-based, Multispecialty Delphi Consensus Document on Stroke Risk Stratification and the Optimal Management of Patients with Asymptomatic and Symptomatic Carotid Stenosis

**DOI:** 10.1101/2025.08.01.25332713

**Authors:** Kosmas I. Paraskevas, Ali F. AbuRahma, Wesley S. Moore, Peter Gloviczki, Bruce A. Perler, Daniel G. Clair, Christopher J. White, Carlo Setacci, Eric A. Secemsky, Peter A. Schneider, Clark J.A.M. Zeebregts, Armando Mansilha, Luca Saba, Ian M. Loftus, Jeffrey Jim, Christos D. Liapis, Vincenzo Di Lazzaro, Alan Dardik, Pavel Poredos, Ankur Thapar, Salvatore T. Scali, Mario D’Oria, Ales Blinc, Alexei Svetlikov, David H. Stone, Sherif A.H. Sultan, Deniz Bulja, Michael C. Stoner, Piotr Myrcha, Maarten Uyttenboogaart, Mark A. Farber, Gianluca Faggioli, Domenica Crupi, Csaba Csobay-Novak, Jens Eldrup-Jorgensen, Gaetano Lanza, Gert J. de Borst, Francesco Stilo, Meghan Dermody, Mauro Silvestrini, Christopher J. Abularrage, Guillaume Goudot, Robert M. Proczka, Gary S. Roubin, Francesco Spinelli, Gabor Menyhei, Saeid H. Shahidi, Jose Ignacio Leal Lorenzo, Arkadiusz Jawien, Tilman Reiff, Laura Capoccia, José Fernandes e Fernandes, Piotr Musiałek, Victor S. Gurevich, Matthew Blecha, Caitlin W. Hicks, Young M. Erben, Mark F. Conrad, Mahmoud B. Malas, Sean P. Lyden, Seemant Chaturvedi, Marc L. Schermerhorn, Andrew N. Nicolaides

## Abstract

**Background:** The optimal management of patients with asymptomatic (AsxCS) and symptomatic (SxCS) carotid stenosis is controversial and includes intensive medical management (i.e., best medical therapy [BMT]) with/without an additional carotid revascularization procedure (i.e., carotid endarterectomy [CEA], transfemoral carotid artery stenting [TFCAS] or TransCarotid Artery Revascularization [TCAR]). The aim of this international, expert-based, multispecialty Delphi Consensus document was to reconcile the conflicting views regarding the optimal management of AsxCS and SxCS patients.

**Methods:** A three-round Delphi Consensus process was performed including 63 experts from Europe (n=37) and the United States (n=26). A total of 6 different clinical scenarios were identified involving patients with either AsxCS or SxCS. For each scenario, 5 treatment options were available: (i) BMT alone, (ii) BMT plus CEA, (iii) BMT plus TFCAS, (iv) BMT plus TCAR, or (v) BMT plus CEA/TFCAS/TCAR. Consensus was achieved when >70% of the Delphi Consensus participants agreed on a therapeutic approach.

**Results:** Most participants concurred that BMT alone is not adequate for the management of a 70-year-old fit male or female patient with 80-99% AsxCS (52/63; 82.5% and 45/63; 71.5%, respectively). In contrast, most panelists would opt for BMT alone for an 80-year-old male AsxCS patient with several co-morbidities (48/63; 76.2%). The majority of participants would opt for BMT plus a carotid revascularization procedure for an 80-year-old male SxCS patient with a recent ipsilateral cerebrovascular event, an ipsilateral 70-99% SxCS and a 5-year predicted risk of ipsilateral ischemic event of 10% (54/63; 85.7%), 15% (59/63; 93.6%), or 20% (63/63; 100%). The opinion of U.S.-based participants varied from that of Europe-based respondents in some scenarios.

**Conclusions:** The present Delphi Consensus document showed that a “one-size-fits-all” approach is not appropriate for patients with either AsxCS or SxCS. Patients should be stratified according to their future stroke risk and should be treated accordingly.

## Introduction

Despite the release of international guidelines by various professional Societies/Organizations (e.g., the Society for Vascular Surgery [SVS],^1,2^ the European Society for Vascular Surgery (ESVS),^3^ the American Heart Association/American Stroke Association^4^ and the European Stroke Organization^5^), there is still substantial controversy regarding the optimal management of patients with asymptomatic (AsxCS) and symptomatic (SxCS) carotid artery stenosis. Treatment options include intensive medical management (i.e., best medical therapy [BMT]) either alone or in combination with a carotid revascularization procedure, namely carotid endarterectomy (CEA), transfemoral carotid artery stenting (TFCAS) or TransCarotid Artery Revascularization (TCAR). Several factors contribute to the ongoing controversy regarding optimal management, including:

a. Variability in physician/surgeon/interventionalist preferences based on individual expertise and/or availability of specific technologies (e.g., TCAR is currently not available in many countries outside the United States);^6^
b. Differences in patient preferences, co-morbidities, anatomical or physiological characteristics and treatment expectations;^7,8^
c. A lack of robust evidence to support strong guidelines recommendations for certain patient subgroups (e.g., women, racial and ethnic minorities, etc.), as these populations were under-represented in landmark randomized controlled trials (RCTs).^3^

As a result, there is often considerable uncertainty about the optimal management of some AsxCS and SxCS patient subgroups. In addition, recent advances suggest that the classification of AsxCS patients based on the degree of carotid stenosis alone may not adequately reflect future stroke risk.^9^

The aim of the present international, multispecialty, expert-based Delphi Consensus document was to address the various therapeutic options available for the management of AsxCS and SxCS patients in an attempt to reconcile the conflicting views.

## Methods

An international, multispecialty, expert-based Delphi consensus document was prepared according to the Conducting and REporting DElphi Studies (CREDES) Checklist.^10^ A total of 26 experts from the United States and 37 experts from Europe (Italy [n=11], Poland [n=4], Netherlands [n=3], United Kingdom [n=2], Russia [n=2], Greece [n=2], Slovenia [n=2], Portugal [n=2], Hungary [n=2], France [n=1], Denmark [n=1], Germany [n=1], Spain [n=1], Bosnia and Herzegovina [n=1], Ireland [n=1] and Cyprus [n=1]) were invited to participate. Overall, 22 of the 26 participants from the U.S. and 25 of the 37 participants from Europe were vascular surgeons. All invited participants had at least 10 years of clinical experience and proof of academic expertise in the management of patients with AsxCS and SxCS, as documented by a list of relevant publications on PubMed/MedLine.

Six different clinical scenarios were identified (**Figure 1**). For each clinical scenario, participants were asked to select the optimal therapeutic approach from the following five options: (a) BMT alone, (b) BMT plus CEA, (c) BMT plus TFCAS, (d) BMT plus TCAR, or (e) BMT plus CEA/TFCAS/TCAR. Option (e) indicated that any revascularization method (CEA, TFCAS or TCAR) could be appropriate for the specific clinical scenario depending on personal expertise and equipment availability.

**Figure 1.**
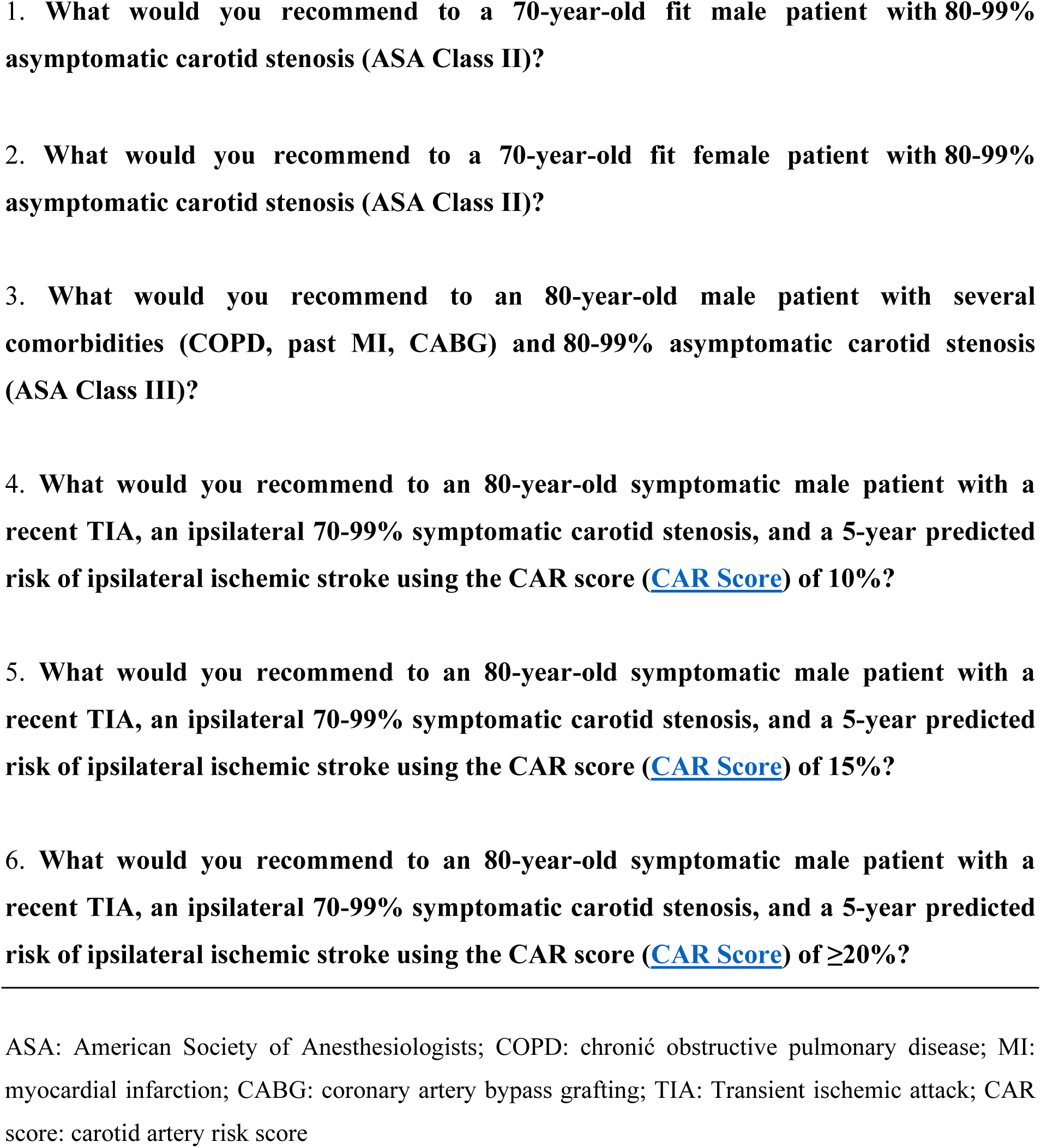
The 6 clinical scenarios included in the Delphi Consensus document

In total, 3 rounds were conducted. Participants had 2 weeks to vote during each round and all voting was anonymous. Only the Delphi Consensus co-ordinator (K.I.P.) had access to individual participant responses. Consensus was defined as >70% agreement among participants on a given therapeutic option. During Round 1, certain issues with the clinical scenarios were identified and clarified. Participants were not informed of the group voting results until after Round 2. In Round 3, participants were asked to finalize their votes.

Differences in responses to each question between U.S. and European participants were assessed using Fisher’s Exact Test. Simulated p-values based on 10,000 Monte Carlo replicates were used due to some cells in the contingency tables containing small or zero counts; Fisher’s test was selected over the chi-squared test to ensure valid inferences under these conditions. To quantify the magnitude and direction of association between geographic region (U.S.A. *vs*. Europe) and treatment preference for AsxCS and SxCS patients, odds ratios (ORs) with 95% confidence intervals (CIs) were calculated with continuity correction applied when necessary to account for zero-cell values. Fisher’s Exact Test was used to calculate p-values.

A pooled analysis of the treatment recommendations for all asymptomatic patient scenarios and all symptomatic patient scenarios in the U.S. respondents versus the European respondents was conducted. There were 26 U.S. respondents and 37 European respondents to 3 asymptomatic and 3 symptomatic scenarios. Therefore, there were a total of 78 U.S. scenario responses versus 111 European scenario responses in both the symptomatic and asymptomatic scenarios. For this pooled analysis the BMT + TCAR response option was merged with the BMT + CEA/CAS/TCAR option as there were very few to no BMT + TCAR responses in several scenarios. Comparison of the frequency of each treatment selection in the U.S. cohort versus the European cohort of respondents was conducted with univariable OR analysis with resultant P-Values, ORs and 95% CIs.

The first draft of the Delphi Consensus document was prepared by the co-ordinator and was circulated to all participants for feedback. The manuscript was revised twice based on their comments and suggestions. The final version of the manuscript was approved by all participants. Any potential conflicts of interest were disclosed and are listed at the end of the manuscript.

## Results

All 63 participants completed all 3 voting rounds. Overall, 22 of 63 (34.9%) maintained the same responses from Round 1 to Round 3, while 18 of 63 (28.6%) changed their votes in at least one scenario between Round 1 and Round 2, but not from Round 2 to Round 3. The remaining 23 of 63 (36.5%) changed their votes to at least one scenario from Round 1 to Round 2 and again from Round 2 to Round 3.

Although consensus on a specific carotid intervention was not achieved, only 11 of 63 (17.4%) participants opted for BMT alone in managing a 70-year-old fit (American Society of Anesthesiologists [ASA] Class II)^11^ male patient with 80-99% AsxCS (**Table 1**). Of these, 4 of 11 (36.3%) would initially choose BMT with plans to re-evaluate the patient with a follow-up ultrasound scan in 6 months to check for disease progression/regression. Furthermore, 7 of 11 (63.7%) would initiate BMT alone while also monitoring for clinical or imaging features suggesting ‘high-stroke’ risk on BMT alone as recommended by the 2023 ESVS guidelines.^3^

**Table 1.** What would you recommend to a 70-year-old fit male patient with 80-99% asymptomatic carotid stenosis (ASA Class II)?

|  | Round 1 | Round 2 | Round 3 |
| --- | --- | --- | --- |
| **BMT alone** | **18 (28.5%)** | **11 (17.4%)** | **11 (17.4%)<sup>671</sup>** |
| **BMT plus CEA** | **25 (39.8%)** | **23 (36.5%)** | **23 (36.5%)<sup>672</sup>** |
| **BMT plus CAS** | **3 (4.8%)** | **2 (3.2%)** | **1 (1.6%)<sup>673</sup>** |
| **BMT plus TCAR** | **–** | **1 (1.6%)** | **–<sup>674</sup>** |
| **BMT plus CEA/CAS/TCAR** | **17 (26.9%)** | **26 (41.3%)** | **28 (44.5%)<sup>675</sup>** |
| **Total** | **63 (100%)** | **63 (100%)** | **63 (100%)<sup>676</sup>** |

Similarly, only 18 of 63 (28.5%) participants selected BMT alone for the management of a 70-yea-old fit (ASA Class II)^11^ female patient with 80-99% AsxCS (**Table 2**). Of these, 8 of 18 (44.4%) opted for an initial BMT alone strategy with reassessment of the patient with a new ultrasound scan in 6 months to check for disease progression/regression. Additionally, 10 of 18 (55.6%) would initiate BMT alone but would also look for clinical/imaging features suggesting ‘high-stroke’ risk on BMT alone, as recommended by the 2023 ESVS guidelines.^3^ In contrast, 48 of 63 (76.2%) panelists would only offer BMT alone to an 80-year-old male patient with 80-99% AsxCS and several comorbidities (ASA Class III), such as chronic obstructive pulmonary disease [COPD], previous myocardial infarction [MI] and/or coronary artery bypass grafting (**Table 3**).

**Table 2.** What would you recommend to a 70-year-old fit female patient with 80-99% asymptomatic carotid stenosis (ASA Class II)?

|  | Round 1 | Round 2 | Round 3 |
| --- | --- | --- | --- |
| **BMT alone** | **28 (44.5%)** | **18 (28.5%)** | **18 (28.5%)** |
| **BMT plus CEA** | **18 (28.5%)** | **19 (30.1%)** | **18 (28.5%)** |
| **BMT plus CAS** | **3 (4.8%)** | **1 (1.6%)** | **2 (3.2%)** |
| **BMT plus TCAR** | – | – | – |
| **BMT plus CEA/CAS/TCAR** | **14 (22.2%)** | **25 (39.8%)** | **25 (39.8%)** |
| **Total** | **63 (100%)** | **63 (100%)** | **63 (100%)** |

**Table 3.**
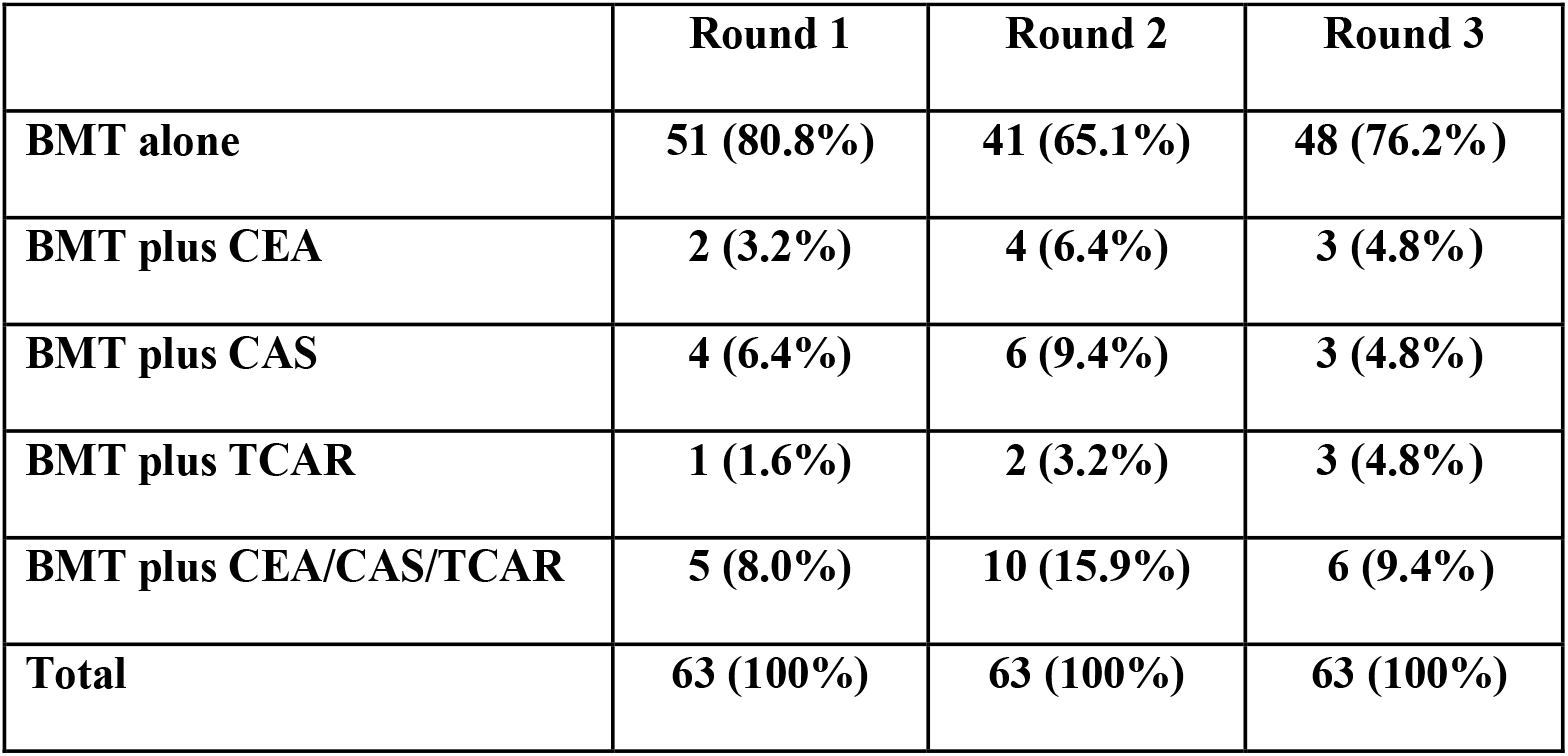
What would you recommend to an 80-year-old male patient with several comorbidities (COPD, past MI, CABG) and 80-99% asymptomatic carotid stenosis (ASA Class III)?

|  | Round 1 | Round 2 | Round 3<sup>20</sup> |
| --- | --- | --- | --- |
| **BMT alone** | **51 (80.8%)** | **41 (65.1%)** | **48 (76.2%)<sup>21</sup>** |
| **BMT plus CEA** | **2 (3.2%)** | **4 (6.4%)** | **3 (4.8%)<sup>22</sup>** |
| **BMT plus CAS** | **4 (6.4%)** | **6 (9.4%)** | **3 (4.8%)<sup>23</sup>** |
| **BMT plus TCAR** | **1 (1.6%)** | **2 (3.2%)** | **3 (4.8%)<sup>24</sup>** |
| **BMT plus CEA/CAS/TCAR** | **5 (8.0%)** | **10 (15.9%)** | **6 (9.4%)<sup>25</sup>** |
| **Total** | **63 (100%)** | **63 (100%)** | **63 (100%)<sup>26</sup>** |

When presented with the clinical scenario involving an 80-year-old symptomatic male patient with a recent transient ischemic attack (TIA) episode, an ipsilateral 70-99% SxCS, and a 5-year predicted risk of ipsilateral ischemic event using the carotid artery risk (CAR) score^12^ of 10%, >80% of participants (54/63; 85.7%) would offer BMT plus an intervention (**Table 4**). This intervention would be CEA or TCAR, but not TFCAS. When the same 80-year-old symptomatic male patient with a recent TIA episode and an ipsilateral 70-99% SxCS, had a 5-year predicted risk of ipsilateral ischemic event using the CAR score^12^ of 15%, >90% of participants (59/63; 93.6%) would offer BMT plus an intervention (**Table 5**), and if the same patient had a CAR score^12^ of ≥20%, all participants (63/63; 100%) would offer BMT plus a carotid intervention (**Table 6**).

**Table 4.**
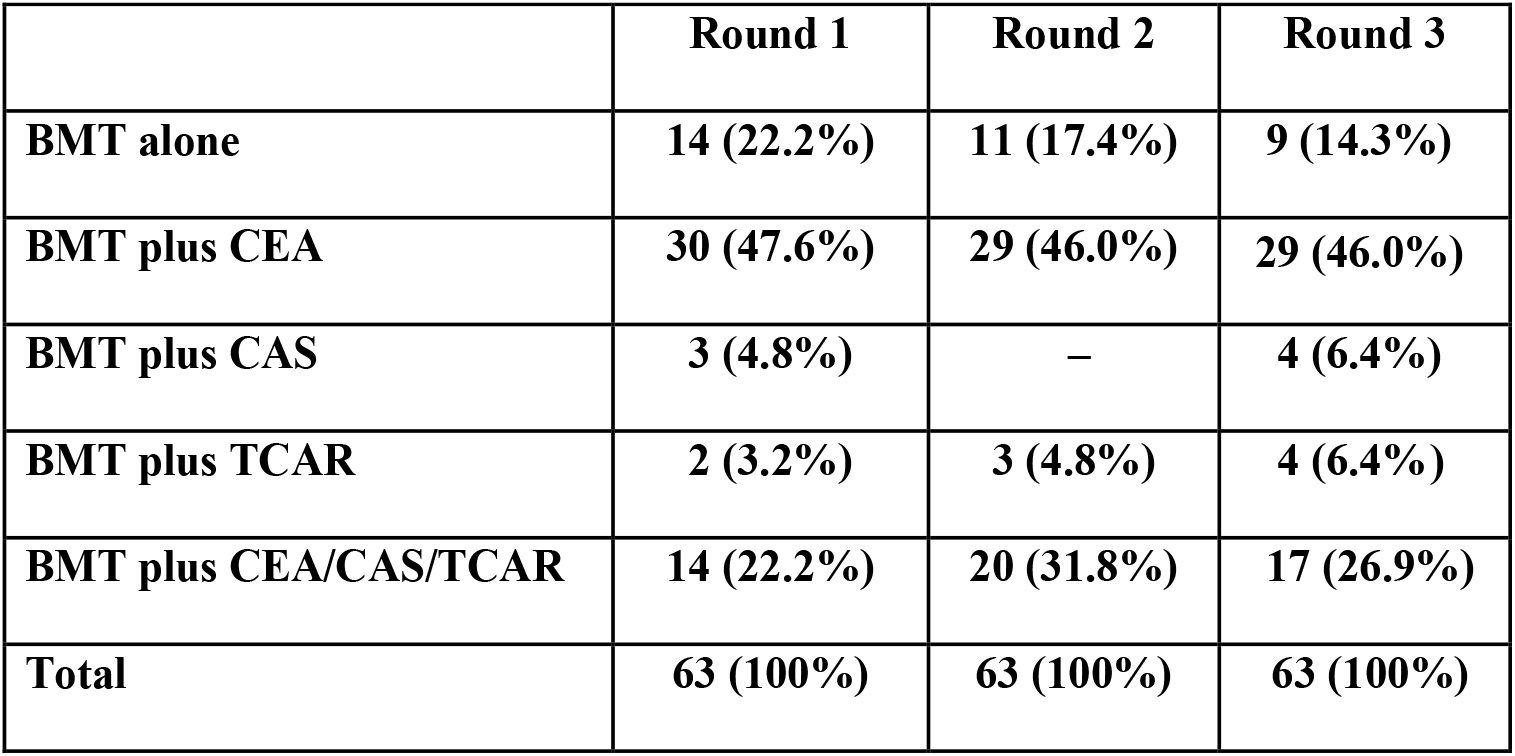
What would you recommend to an 80-year-old symptomatic male patient with a recent TIA, an ipsilateral 70-99% symptomatic carotid stenosis, and a 5-year predicted risk of ipsilateral ischemic stroke using the CAR score (CAR Score) of 10%?

|  | Round 1 | Round 2 | Round 3 |
| --- | --- | --- | --- |
| **BMT alone** | **14 (22.2%)** | **11 (17.4%)** | **9 (14.3%)<sup>746</sup>** |
| **BMT plus CEA** | **30 (47.6%)** | **29 (46.0%)** | **29 (46.0%)<sup>747</sup>** |
| **BMT plus CAS** | **3 (4.8%)** | **–** | **4 (6.4%)<sup>748</sup>** |
| **BMT plus TCAR** | **2 (3.2%)** | **3 (4.8%)** | **4 (6.4%)<sup>749</sup>** |
| **BMT plus CEA/CAS/TCAR** | **14 (22.2%)** | **20 (31.8%)** | **17 (26.9%)<sup>750</sup>** |
| **Total** | **63 (100%)** | **63 (100%)** | **63 (100%)<sup>751</sup>** |

**Table 5.** What would you recommend to an 80-year-old symptomatic male patient with a recent TIA, an ipsilateral 70-99% symptomatic carotid stenosis, and a 5-year predicted risk of ipsilateral ischemic stroke using the CAR score (CAR Score) of 15%?

|  | Round 1 | Round 2 | Round 3 |
| --- | --- | --- | --- |
| **BMT alone** | **4 (6.4%)** | **6 (9.5%)** | **4 (6.4%)<sup>1</sup>** |
| **BMT plus CEA** | **30 (47.5%)** | **32 (50.8%)** | **29 (46.0%)<sup>2</sup>** |
| **BMT plus CAS** | **4 (6.4%)** | **1 (1.6%)** | **3 (4.8%)<sup>3</sup>** |
| **BMT plus TCAR** | **4 (6.4%)** | **2 (3.2%)** | **3 (4.8%)<sup>4</sup>** |
| **BMT plus CEA/CAS/TCAR** | **21 (33.3%)** | **22 (34.9%)** | **24 (38.0%)<sup>5</sup>** |
| **Total** | **63 (100%)** | **63 (100%)** | **63 (100%)<sup>6</sup>** |

**Table 6.** What would you recommend to an 80-year-old symptomatic male patient with a recent TIA, an ipsilateral 70-99% symptomatic carotid stenosis, and a 5-year predicted risk of ipsilateral ischemic stroke using the CAR score (CAR Score) of ≥20%?

|  | Round 1 | Round 2 | Round 3 |
| --- | --- | --- | --- |
| **BMT alone** | – | **1 (1.6%)** | – 796 |
| **BMT plus CEA** | **31 (49.4%)** | **29 (46.0%)** | **30 (47.5%)<sup>797</sup>** |
| **BMT plus CAS** | **6 (9.3%)** | **3 (4.8%)** | **4 (6.4%)<sup>798</sup>** |
| **BMT plus TCAR** | **4 (6.4%)** | **2 (3.2%)** | **3 (4.8%)<sup>799</sup>** |
| **BMT plus CEA/CAS/TCAR** | **22 (34.9%)** | **28 (44.4%)** | **26 (41.3%)<sup>800</sup>** |
| **Total** | **63 (100%)** | **63 (100%)** | **63 (100%)<sup>801</sup>** |

Comparative analysis between U.S.-based and Europe-based participants revealed some regional differences (**Supplementary Tables 1-6**). European participants were more likely to opt for BMT alone in the management of a 70-year-old, fit male (21.6% *vs*. 11.5%, respectively; **Supplementary Table 1**) and female (35.1% *vs*. 19.2%, respectively; **Supplementary Table 2**) patients compared with their North American counterparts, but these differences were not significant. Most U.S. and European participants opted for BMT alone for the management of an 80-year-old male patient with several comorbidities (COPD, past MI, CABG) and 80-99% AsxCS (73.4% vs. 78.4%, respectively; p=0.3688; **Supplementary Table 3**). European participants favored BMT plus CEA as the optimal treatment option for an 80-year-old symptomatic male patient with a recent TIA, an ipsilateral 70-99% SxCS, and a 5-year predicted risk of ipsilateral ischemic stroke using the CAR score of 10% (48.6% *vs*. 42.4%, respectively; p=0.119), 15% (56.8% *vs*. 30.8%, respectively; p=0.0124) and 20% (59.5% *vs*. 30.8%, respectively; p=0.0032) compared with U.S. participants. In contrast, U.S. participants were more likely to offer CEA or TCAR (but not TFCAS) to these patients compared with European participants (42.4% *vs*. 16.3%, 61.6% *vs*. 21.6% and 65.4% *vs*. 14.3%, respectively; **Supplementary Tables 4, 5 and 6**).

The three asymptomatic patient scenarios were pooled to allow for comparison of U.S. *vs*. European participant preferences. U.S.-based participants were significantly more likely than their European counterparts to opt for BMT plus CEA/CAS/TCAR (42.3 *vs*. 26.0%, respectively; OR: 4.251; 95% CI: 2.204-8.364, p<0.001; **Supplementary Table 7**). The BMT + TCAR option was merged into the BMT + CEA/TFCAS/TCAR cohort for this analysis due to low sample size on the BMT + TCAR option and the current minimal availability of TCAR in Europe.

The three symptomatic patient scenarios were pooled to allow for comparison of U.S.-based *vs*. European participant preferences. U.S.-based participants were significantly more likely to opt for BMT plus CEA/CAS/TCAR in symptomatic patients (60.3% *vs*. 27.0%, respectively; OR: 4.060; 95% CI: 2.112-7.958; p<0.001). In contrast, U.S.-based participants were significantly less likely to opt for BMT + CEA or BMT + CAS than their European counterparts in treating SxCS (34.6 *vs*. 54.9%, respectively; OR: 0.436; 95% CI: 0.228-0.822; p=0.008; and 0 *vs*. 10.0%, respectively; OR: 0; 95% CI: 0-0.533; p=0.003; **Supplementary Table 8**). Once again, the BMT + TCAR option was merged into the BMT + CEA/TFCAS/TCAR cohort for this analysis due to low sample size on the BMT + TCAR option and the current minimal availability of TCAR in Europe.

## Discussion

The present international, expert-based Delphi Consensus document revealed several findings regarding the optimal management of both AsxCS and SxCS patients. These findings are presented and discussed.

### AsxCS patients

It is now well-recognized that not all AsxCS patients carry the same stroke risk. A 2014 opinion article emphasized the importance of identifying ‘high-risk’ AsxCS patients to selectively offer prophylactic carotid revascularization procedures to those most likely to benefit.^14^ A number of clinical and imaging features have been proposed for stratifying stroke risk in this population, including: (1) the detection of microemboli on transcranial Doppler, identification of the unstable carotid plaque using ultrasound, (3) reduced cerebrovascular reserve, (4) identification of intraplaque hemorrhage on MRI, (5) progression in the stenosis severity, and, (6) a combination of multiple independent risk stratification parameters (e.g., baseline degree of stenosis, history of contralateral stroke or TIA, size of juxtaluminal plaque area ≥8 mm^2^ without a visible echogenic cap and the presence of discrete white areas in a hypoechoic plaque, or a combination of a low gray scale median score with transcranial Doppler microembolic signals).^14^ Based on these findings, the 2017 ESVS carotid guidelines recommended that for patients with 60-99% AsxCS and 1 or more of these ‘high-risk’ clinical/imaging features associated with an increased risk for late stroke on BMT alone,^14^

CEA should be considered (Class IIa; Level of Evidence: B) or TFCAS may be considered (Class IIb; Level of Evidence: B) for the reduction of long-term risk of stroke, provided that anatomy is favorable, 30-day stroke/death rates are ≤3% and the patient’s life-expectancy exceeds 5 years.^15^ These recommendations remained unchanged in the recently updated 2023 ESVS carotid guidelines.^3^

Similarly, the 2022 SVS carotid guidelines endorse the use of CEA, TCAR or TFCAS in patients with ≥70% AsxCS, provided that the patient has at least a 3-to 5-year life expectancy and perioperative stroke/death rates can be ≤3%.^1,2^ The SVS Guidelines emphasized that selection of the revascularization strategy should be based on the presence or absence of specific high-risk anatomic criteria for each procedure.^1,2^ For instance, the presence of a tracheal stoma or a lesion above C2 would be a contraindication for CEA.^1,2^ In contrast, a distance to the carotid bifurcation <5 cm or a common carotid artery diameter <6 mm would be a contraindication for TCAR.^1,2^ Finally, a tortuous common or internal carotid artery would be a contraindication for TFCAS.^1,2^ In addition, the SVS carotid guidelines clearly indicated that lesion morphology such as echolucency, calcification, long irregular plaques, the presence of fresh thrombus or a string sign can affect outcomes and may alter decision-making concerning the optimal carotid revascularization procedure.^1,2^ Therefore, the choice of the optimal therapeutic modality would depend on the presence or absence of such high-risk anatomic criteria and lesion morphology.^1,2^ Finally, it was specified that physiologic comorbidities such as congestive heart failure, left ventricular ejection fraction ≤35%, unstable angina, the presence of MI within the past 6 weeks COPD and renal failure constitute considerable physiologic risks, and in such patients, TCAR is preferred over CEA and TFCAS.^2^

Recent evidence suggests that the degree/percentage of AsxCS alone is not an adequate predictor of future ipsilateral ischemic stroke risk. Several other parameters have been proposed to more accurately stratify AsxCS patients with regards to future stroke risk. Examples include the type of carotid plaque,^13^ or a high carotid plaque-reporting and data system score (RADS).^9^ These parameters may be more accurate predictors of future ipsilateral ischemic stroke risk and should probably be implemented in future guidelines to guide the identification of ‘high-risk’ AsxCS individuals for whom a prophylactic carotid intervention is warranted in addition to BMT. Identification of prognostic factors for long-term survival in AsxCS patients and risk prediction models for the development of a future stroke in these individuals, as well as the development of valid and reliable stroke risk stratification models/systems are crucial to select those asymptomatic patient subgroups most likely to benefit from a prophylactic carotid intervention.^16–20^

### SxCS patients

Both the 2022 SVS and the 2023 ESVS carotid guidelines strongly recommend carotid revascularization within 14 days of an ischemic cerebrovascular in patients with and an ipsilateral 50-99% SxCS.^1–3^ In this scenario, both guidelines advocate for CEA over TFCAS, based on evidence supporting superior safety and efficacy in these patients.^1–3^ Additionally, the SVS guidelines highlight data from large national registries suggesting that in symptomatic patients, TCAR is associated with lower stroke/death rates than TFCAS and demonstrates comparable outcomes with CEA.^2^ However, it is important to note that the vast majority of TCAR procedures to date have been performed in patients considered high risk for CEA due to anatomic or medical factors. While early results are encouraging, further data in low-risk symptomatic patients are needed to validate these findings.^2^

A few months ago, Dermody (Penn Medicine Lancaster, U.S.A.) presented at VIVA 2024 Meeting in Las Vegas the early data on TCAR from the ROADSTER 3 trial.^21^ This is an FDA post-approval study of the first prospective multicenter trial evaluating the safety and efficacy of TCAR using the Silk Road Enroute transcarotid stent system in conjuction with its Enroute transcarotid neuroprotection system for the treatment of carotid stenosis in standard surgical risk patients.^21^ ROADSTER 3 enrolled 344 patients at 53 U.S. sites for intention-to-treat (ITT) analyses. The single arm study primary end-point was a composite of major adverse events of stroke, death and MI through 30 days post-procedure and ipsiliateral strokes from day 31 to 365 post-procedure. Approximately 75% of patients were younger than 75 years of age, 43% were female and 16% had SxCS. The mean lesion length was 23 mm and 17% of the lesions had severe calcification. The 30-day rate of stroke/death/MI in the study’s ITT population was 0.9%, which decreased to 0.6% within per-protocol analysis involving 320 patients. The incidence of cranial nerve injury within 30 days was 0.6% and all cranial nerve deficits resolved within 6 months.^21^ It was concluded that since TCAR is less invasive than CEA and has similar stroke rates with a lower incidence of cranial nerve injury, if a patient is able to take dual antiplatelet and statin therapy and has anatomy that is amenable to TCAR, then TCAR should be the first-choice modality.^21^

Recently, the 2-year interim results of the 2nd European Carotid Surgery Trial (ECST-2) were published.^22^ ECST-2 was an international, multicenter (n=30), open-label, non-inferiority randomized controlled trial conducted at sites in Europe and Canada.^22^ The trial tested whether patients with ≥50% AsxCS or SxCS with a low-to-intermediate predicted risk of stroke receiving BMT would benefit from additional revascularization.^22^ The primary end-point for the 2-year analysis was a ‘hierarchical’ composite outcome consisting of: (1) periprocedural death, fatal stroke, or fatal MI; (2) non-fatal stroke; (3) non-fatal MI; or (4) new silent cerebral infarction on imaging.^22^ The interim analysis found no significant difference in the ‘hierarchical’ outcome between treatment groups.^22^ It was therefore concluded that ‘***no evidence for a benefit of revascularization in addition to BMT was found in the first 2 years following treatment for patients with asymptomatic or symptomatic carotid stenosis of 50% or greater with a low or intermediate predicted stroke risk (assessed by the CAR score)’***.^22^ They further recommended that ‘***patients with asymptomatic and low or intermediate risk symptomatic carotid stenosis should be treated with BMT alone until further data from the 5-year analysis of ECST-2 and other trials become available’***.^22^

Although ECST-2 was an exemplary RCT, its findings raise some concerns. Notably, the composite outcome of periprocedural death, stroke, or MI, was 10.2% in the BMT alone group compared to 10.5% in the BMT plus revascularization group (hazard ratio: 0.92; 95% CI: 0.51-1.67; p=0.46).^22^ Given that the 2-year MI rate in the BMT plus revascularization group was 2.5%, this implies a periprocedural death/stroke rate of ≥8.0%.^22^ Both the 2022 SVS^1,2^ and the 2023 ESVS^3^ clearly state the periprocedural stroke and death rates should be ≤3% in AsxCS and ≤6% in SxCS patients to justify revascularization. Accordingly, the periprocedural death, stroke or MI rates reported for the BMT plus carotid revascularization group in ECST-2^22^ are alarmingly high and exceed the recommended safety thresholds endorsed by both the SVS and the ESVS guidelines, for patients with either AsxCS or SxCS undergoing a carotid intervention.

ECST-2^22^ had several flaws/limitations:

1. Although a sample size of 2,000 patients was initially calculated,^23^ at the end only 428 patients were randomized to BMT alone (n=214) or BMT plus revascularization (n=214),^22^ which is an extremely small sample to predict major cerebrovascular events.
2. Both symptomatic (40%) and asymptomatic (60%) patients were included, and even within the symptomatic patients, only those at low/intermediate risk based on CAR scoring were considered.
3. Among the asymptomatic patients, 47 of 129 (36.5%) in the BMT group and 48 of 129 (37.2%) in the BMT plus revascularization group had AsxCS ≤69%.^22^ It would be interesting to know why nearly 1 for every 4 asymptomatic patients in the BMT plus revascularization group (48 of 214; 22.5%) was offered an intervention for a ≤69% AsxCS.^22^
4. Among the 214 patients allocated to BMT alone, 22 (10%) eventually underwent an ipsilateral carotid revascularization procedure.^22^

These flaws/limitations suggest that long-term data and a bigger sample size is needed before we consider the conclusions of this study.

Although guidelines provide recommendations for patients at ‘average surgical risk’, the management may differ in those deemed ‘high-risk’ due to medical comorbidities. In AsxCS patients, for example, life expectancy of >5 years and low perioperative complications are prerequisites to reliably achieving benefit from a carotid intervention.^1–3^ Furthermore, a systematic review of outcomes in 21 registries (>1,500,000 patients) showed that stroke and death rates after TFCAS often exceed the recommended thresholds by the AHA/ASA (<3% for AsxCS; <6% for SxCS).^24^ More nuanced stroke risk stratification tools such as the carotid plaque-RADS classification system^9^ or various plaque features (e.g., ulceration, thin fibrous cap, juxtaluminal black area >8mm^2^),^13^ and patient-specific anatomical and physiological characteristics may help identify which individuals would benefit most from either BMT alone or BMT plus carotid revascularization.

The differences between participants from the U.S. and their European counterparts in the pooled analysis probably reflect the availability and increased utilization of TCAR in the United States. TCAR offers a less invasive carotid revascularization option than CEA with similar perioperative stroke rates when applied to patients with anatomy appropriate for TCAR.^25^ TCAR is therefore viewed in the U.S. as a viable and potentially superior option to CAS and CEA in patients at higher surgical risk due to medical comorbidities or due to anatomically hostile necks.

The present Delphi Consensus document has some limitations. As with all Delphi Consensus documents, a different composition of the panel (e.g., more interventional radiologists/cardiologists, fewer vascular surgeons, more U.S. participants, etc.) could have produced different results. In addition, the decision-making for each patient would also take into account individual anatomical and physiological characteristics/risk factors that would make them high-risk for specific procedures (e.g., anatomy suitable for the selected procedure, patient life expectancy, type of carotid plaque, etc.). Furthermore, no information was provided regarding the type of plaque (e.g., the presence of carotid ulcer, a thin fibrous cap, discrete white areas or large a juxtaluminal black area [>8 mm^2^]) or the brain CT findings (e.g., the presence of silent ipsilateral infarcts). For example, there is evidence that intraplaque hemorrhage and plaque ulceration are more likely in patients with mild-to-moderate SxCS than in high-grade AsxCS/SxCS.^26^ Such information could alter the votes of many of the participants. Finally, the Carotid Revascularization and Medical Management for Asymptomatic Carotid Stenosis (CREST-2)^27^ study is on-going. Its results may alter the opinion of some of the participants of this Delphi Consensus.

## Conclusions

In conclusion, this international, multi-specialty Delphi Consensus Document highlights the ongoing variability in the management of both SxCS and AsxCS patients. Although consensus was not achieved in all scenarios – particularly regarding the preferred revascularization technique – these differences largely reflect the diverse expertise, geographic practice patterns, and resource availability among panel participants.

Notably, the panel agreed that BMT alone is insufficient for most patients with SxCS, and that select subgroups of AsxCS patients may also benefit from revascularization, especially when high-risk features are present. These findings support the importance of personalized stroke risk stratification, incorporating clinical, anatomical, and imaging features to guide decisions about whether and how to intervene.

As emerging data – such as from ECST-2^22^ and CREST-2^27^ – continue to shape the evidence base, interdisciplinary dialogue and individualized decision-making will remain critical in optimizing outcomes for patients with both AsxCS and SxCS.

## Data Availability

All data is available by Dr. Kosmas I. Paraskevas

## Acknowledgements

The authors would like to thank Dr. Camila R. Guetter, MD, MPH from the Department of Surgery, Beth Israel Deaconess Medical Center, Boston, U.S.A., for her valuable help with the statistics of this manuscript.

